# Global Assessment of the Relationship between Government Response Measures and COVID-19 Deaths

**DOI:** 10.1101/2020.07.04.20145334

**Authors:** Thomas Hale, Andrew J. Hale, Beatriz Kira, Anna Petherick, Toby Phillips, Devi Sridhar, Robin N. Thompson, Samuel Webster, Noam Angrist

**Affiliations:** Blavatnik School of Government, University of Oxford; Larner College of Medicine at the University of Vermont; Professor, University of Edinburgh; Mathematics Institute, University of Oxford; Unaffiliated

## Abstract

**Objective:** To provide an early global assessment of the impact of government stringency measures on the rate of growth in deaths from COVID-19. We hypothesized that the overall stringency of a government’s interventions and the speed of implementation would affect the growth and level of deaths related to COVID-19 in that country.

**Design:** Observational study based on an original database of global governmental responses to the COVID-19 pandemic. Daily data was collected on a range of containment and closure policies for 170 countries from January 1, 2020 until May 27, 2020 by a team of researchers at Oxford University, UK. These data were combined into an aggregate stringency index (SI) score for each country on each day (range: 0-100). Regression was used to show correlations between the speed and strength of government stringency and deaths related to COVID-19 with a number of controls for time and country-specific demographic, health system, and economic characteristics.

**Interventions:** Nine non-pharmaceutical interventions such as school and work closures, restrictions on international and domestic travel, public gathering bans, public information campaigns, as well as testing and contact tracing policies.

**Main outcomes measures:** The primary outcome was deaths related to COVID-19, measured both in terms of maximum daily deaths and growth rate of daily deaths.

**Results:** For each day of delay to reach an SI 40, the average daily growth rate in deaths was 0.087 percentage points higher (0.056 to 0.118, P<0.001). In turn, each additional point on the SI was associated with a 0.080 percentage point lower average daily growth rate (−0.121 to −0.039, P<.001). These daily differences in growth rates lead to large cumulative differences in total deaths. For example, a week delay in enacting policy measures to SI 40 would lead to 1.7 times as many deaths overall.

**Conclusions:** A lower degree of government stringency and slower response times were associated with more deaths from COVID-19. These findings highlight the importance of non-pharmaceutical responses to COVID-19 as more robust testing, treatment, and vaccination measures are developed.

## Introduction

The current pandemic of SARS-CoV-2 and resultant COVID-19 disease have upended healthcare, cultural, financial, and government systems worldwide. As of June 1, 2020, there have been over 6 million confirmed cases and over 371,000 deaths reported in over 200 countries and territories.^1^ There are currently no approved vaccines available,^2–4^ and only recently was an antiviral agent approved, meaning that control of the COVID-19 pandemic has largely relied on increasingly stringent government non-pharmaceutical interventions (NPIs).^5–10^

As the outbreak has progressed, such interventions have proliferated worldwide (see Figure 1 in supplement), including school closings, travel restrictions, public gathering bans, and stay-at-home orders. Such policies aim to create physical distancing or otherwise slow the spread of COVID-19, often in concert with testing and contact tracing regimes of varying robustness.^7,11–14^ In some cases, closure and containment measures have been extreme, with unprecedented social, cultural, and financial implications.^15–17^ Governments have varied significantly in both the degree of their interventions and how quickly they adopt them.^18,19^ The efficacy of specific interventions on disease-prevention is only now being rigorously assessed, and more data are needed.^14,19–26^

**Figure 1:**
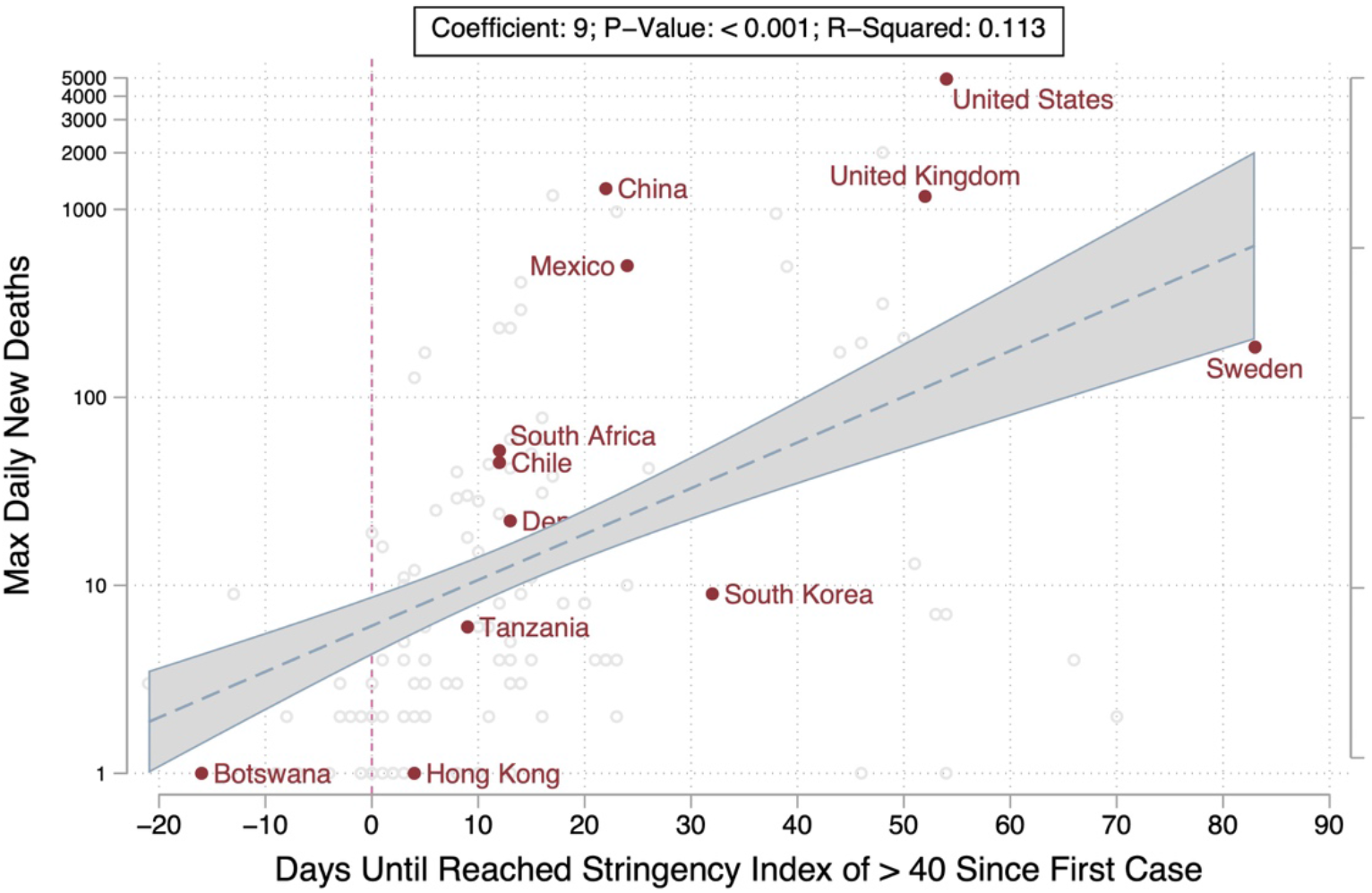
Speed of Government Response and Peak Deaths Each additional day delayed to reach a government stringency index score of 40 (on a scale of 0 to 100) since the first case in a given country is associated with 9 (95% CI 4.9-13.2) more daily deaths at the peak of the epidemic curve. The dotted zero line on the x-axis is the time between first case in a country and when the government stringency index reached 40. Values greater than the 0 line indicate that a country reached a response index of 40 after the first case in that country. Values below the zero line indicate a government response of 40 was reached before the first in-country case. This threshold was chosen since it correlated with a series of assertive policies such as stay-at-home orders, cancelling public events, closing work, among others, and was eventually reached by nearly all countries. The estimates shown are produced on a non-log scale, and the y-axis is shown on a log scale for ease of visual representation.

We present a global analysis of governments’ responses to date, and a first global assessment of their relationship to the spread of the pandemic. We tracked 170 governments’ responses across a series of non-pharmaceutical interventions and created a composite index that captured how, over time, each country’s government responded. We believe this dataset is the most comprehensive view of governmental response to COVID-19 yet created, presenting cross-temporal and cross-country data since January 1, 2020. We hypothesized that the overall stringency of governments’ interventions and the speed of their implementation would affect the rate of deaths related to COVID-19, providing an empirical test of a common assumption in epidemiological models.

## Methods

### Data collection

We collected information on 170 national governments’ responses across a range of NPIs (see Table S1 in supplement). These measures were recorded for each day in each country, creating a measure of variation in government responses both across countries and across time. Data were collected by the authors and trained research assistants from publicly available sources such as news articles, government briefings, and international organizations. Data collectors coded government responses on a simple binary or ordinal scale registering the stringency of a given policy. Several indicators were further classified as either “targeted” (meaning they apply only in a geographically concentrated area) or “general” (meaning they apply throughout the entire jurisdiction). The data cover over 170 countries from January 1, 2020, through May 27, 2020, though not all of the analyses below include all countries due to limitations in some of the supplementary data sources described below. Because we do not use human data or tissue, or involve human subjects, approval by the university review board was not required. To ensure accuracy and consistency, data collectors were required to pass an online training and to participate in regular team review meetings. Each data point was verified by at least two data collectors independently, and includes notes and source materials to substantiate each observation. Importantly, for all NPIs, we record only the official policies at the national level, not how well they are implemented or enforced.

### Measuring the speed and degree of government non-pharmaceutical interventions

Our primary measure of governments’ NPIs is a composite Stringency Index (SI) that records the number and restrictiveness of government containment and closure measures, calculated as follows. For each of the nine relevant policy response indicators, we create a score by taking the ordinal value, adding 0.5 if the policy is general rather than targeted, and rescaling each of these to range from 0-100. Conservatively, we assign a score of 0 to any indicators missing data, and reject any country-days where more than one of the indicators is missing. The mean of these nine scores gives the composite SI.

We rely chiefly on this simple, unweighted SI because this approach is most transparent and easiest to interpret.^27^ Composite indices have the value of facilitating comparison across countries, albeit with the trade-off of condensing information. In practice, however, we observe that most countries in the time period of analysis adopted most NPIs as a package, further justifying the use of a composite measure (Figure S1 in supplement). As a robustness check, we used principal component analysis and principal factor analysis to examine whether the indicators that comprise the index can be treated as capturing a single dimension.^28^ Principal factor analysis (Table S2 in supplement) showed the eigenvalue for the first estimated factor accounts for 84 percent of the total variance, strongly supporting this approach.

The degree of government response is measured by the value of the SI for a country on a particular day, or, in the cross-sectional analyses, the average level of stringency from January 1, 2020 to May 27, 2020. The speed of government response is calculated as the number of days between when a country records its first COVID-19 case and when it reaches a stringency level of 40 out of 100. This threshold was chosen because, observationally, we see that almost every country reaches at least this threshold at some point, but countries vary significantly in when they reach this mark. While the majority of countries eventually exceed 80 at their most stringent point, a threshold of 40 allows us to capture the point at which all countries but four start a substantive response.

In addition to the nine indicators that comprise the stringency index, we include as a control measures of governments’ testing and contact tracing (Table S1 in supplement). These are also recorded on an ordinal scale representing the breadth and thoroughness of the policy.

### Outcome variables

We sought to estimate the relationship between government interventions and the intensity of the COVID-19 outbreak by country. Information on confirmed COVID-19 cases and deaths were taken from the European Centre for Disease Prevention and Control, supplemented with data from Johns Hopkins University for the autonomous Chinese regions of Hong Kong and Macau.^29,30^ However, the true number of cases and deaths, as well as the reproduction number, are the subjects of significant uncertainty and a major topic of ongoing research.^15,31–33^ Observationally, the true number of cases is difficult to measure consistently because different countries have tested for COVID-19 more or less widely, and report case information in variable ways.^7,34^ We take a conservative approach and use recorded deaths as our main outcome, which we expect to be reported more consistently and captures the public health consequence of the epidemic most directly.

We use two measures of deaths to capture both the “slope” and “peak” (to the extent it yet exists) of the epidemiological curve. First, we analyse growth in deaths in terms of the daily log difference in deaths in a country. We use the log transformation to account for the exponential trajectory of the epidemic growth curve. For cross-sectional analyses, we also look zat the average growth rate since first case in each country until May 27, 2020. Second, we consider the maximum daily number of new deaths, that is the “peak” in the number of deaths.

### Statistical analysis

Our approach aligns with observational methods to study how government measures address historical epidemics.^35^ We estimate how the speed and degree of government response relates to growth in deaths and the maximum number of new daily deaths using Ordinary Least Squares (OLS) regression. We estimate both cross-sectional models in which countries are the unit of analysis, as well as longitudinal models on time-series panel data with country-day as the unit of analysis. In the latter, we estimate models that use both time and country fixed effects. The former shows the conditional effect of government responses at a given day in the course of the outbreak across countries; the latter shows the conditional effect of changes in government stringency within countries over time.^36^ Importantly, country fixed effects control for all country-specific characteristics that do not vary over the period of analysis, such as the level of wealth, pre-existing robustness of the health care system, the government’s overall capacity to implement policy, or the population’s general tendency to follow government advice or not. We also estimate models that directly include data from the World Bank on the dependency ratio (to capture the age structure of a given country), population density, health capacity in terms of beds and physicians per capita, and GDP per capita.

This kind of observational study faces two key inferential challenges: how to control for the effect of time, including the “natural” growth and diminishing of the disease, which is unobserved, and the possibility of reverse causality or other forms of endogeneity.^37^ For example, a positive relationship between current stringency and new deaths does not necessarily mean stringency increases deaths; rather deaths might trigger a policy response. We address these issues in three ways. First, in the time series models we lag the explanatory variable by six weeks, looking at the effect of past stringency on current changes in the outcome.^38^ Second, in the time series analyses, we use the difference in logs as the dependent variable, approximating the shape of an epidemiological curve. Third, we include a time term - the number of days since the first case - in regressions to account for natural growth patterns in deaths as well as normalize the time period to account for the time in which the epidemic reached different countries across the globe. All analyses were performed using Stata version 15.01. We cluster all standard errors at the country level to account for autocorrelation by day within a country.

## Results

### Variation in the speed and intensity of government responses

We observed significant variation in both the level of stringency and the time at which policies are adopted across national governments. While there is a near-universal increase in countries adopting containment and closure measures over time, with most countries moving to stringent measures after the first week of March 2020, the varied spread of the disease globally means that some countries adopted “lockdown” measures before local transmission began and some after. Countries tended to follow a common sequence (Figure S1 in supplement).

Our core question is whether variation in speed and degree of response can be associated with changes in the maximum number of daily deaths or the rate of growth in deaths. Figures 1–4 explore these relationships. Figure 1 shows a positive correlation between the maximum number of daily deaths and a delayed response measured in days since the first case. Countries that delayed action, such as the United States, the United Kingdom, or Sweden, see a higher total level of deaths than places like South Korea, Hong Kong, or Botswana, with each day of delay in reaching SI 40 corresponding with 9 (95% CI 4.9-13.2) additional deaths on the peak day.

Figure 2 instead looks at the growth rate in deaths since first case to May 27, 2020, showing the same positive correlation between slower responses and higher average daily growth rates in deaths. Specifically, each additional day between a country’s first case and reaching SI 40 is observed to correlate with an average daily growth rate 0.083 percentage points higher. In other words, a country that acted a month later than another country would see an average daily growth rate 2.49 percentage points higher. Because even small changes in daily growth rates can over time translate into large differences in total deaths, the observed correlation indicates starkly different outcomes.

**Figure 2:**
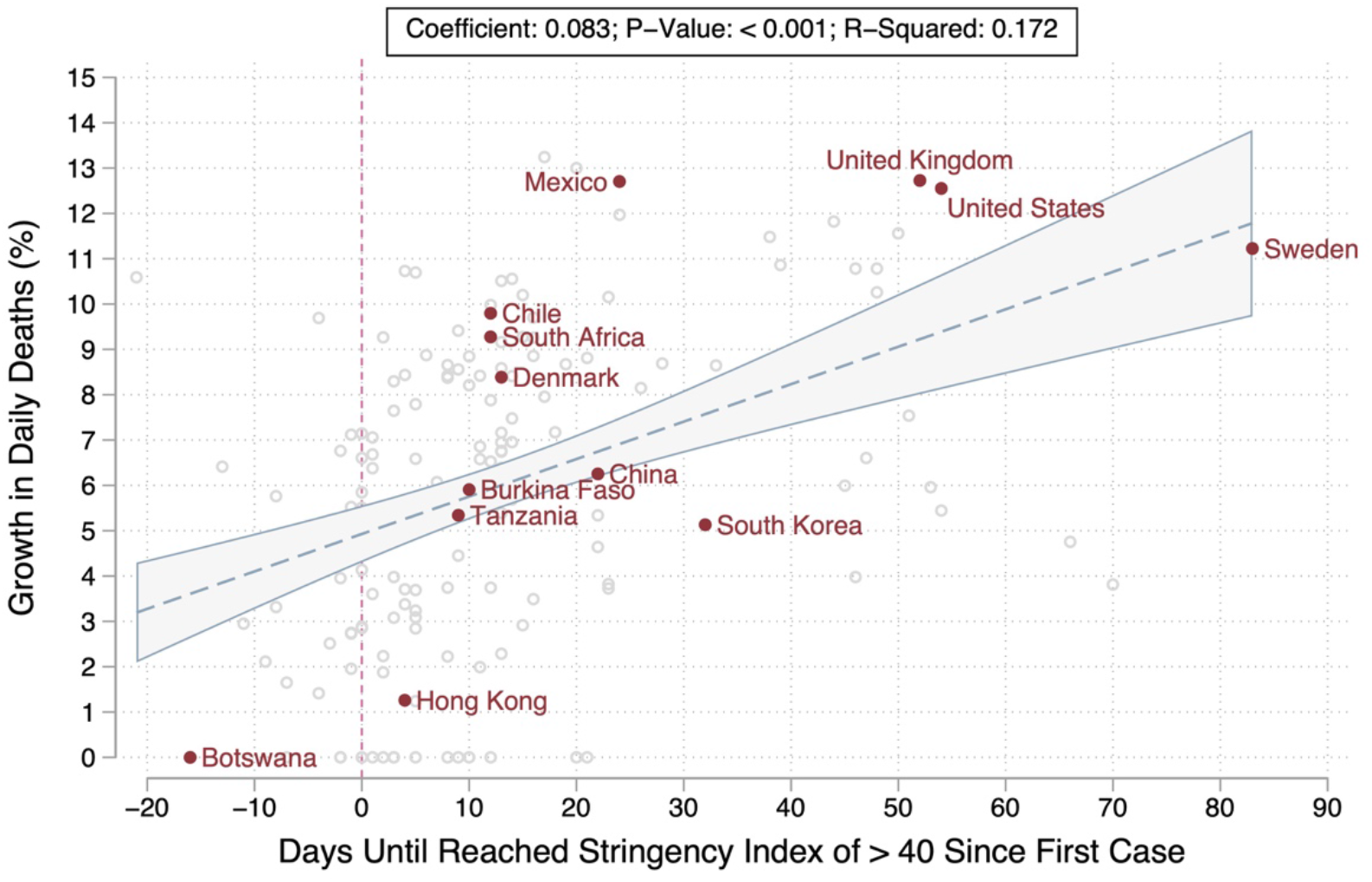
Speed of Government Response and Growth in New Deaths Each additional day delayed to reach a government stringency index score of 40 (on a scale of 0 to 100) is associated with an 0.083 (95% CI 0.054-0.111) percentage point higher growth rate in daily deaths. That is, a country that acted 30 days later than another country would see an average daily growth rate approximately 2.5 percentage points higher. The dotted zero line on the x-axis is the time between first case and when the government reached SI 40. Values greater than the 0 line indicate that a country reached a response index of 40 after the first case. Values below the zero line indicate a government response of 40 was reached before first case. This threshold was chosen since it correlated with a series of assertive policies such as stay-at-home orders, cancelling or public events, closing work, among others, and was reached by nearly all countries but varied in timing of when it was reached. We average growth in deaths since the time of first case for each country to make comparisons on an equivalent time scale. Growth rates are computed as the first differences on a log scale.

Figure 3 considers not speed but degree of government response, showing a negative relationship between countries’ average score on the stringency index since the date of their first case, and the average rate of growth in deaths six weeks later. Each additional point of stringency of government response is associated with a 0.072 percentage point reduction in the growth rate of daily deaths. That is, a country that had the maximum score (100) would see an average daily growth rate 7.2 percentage points lower than a country with no measures in place.

**Figure 3:**
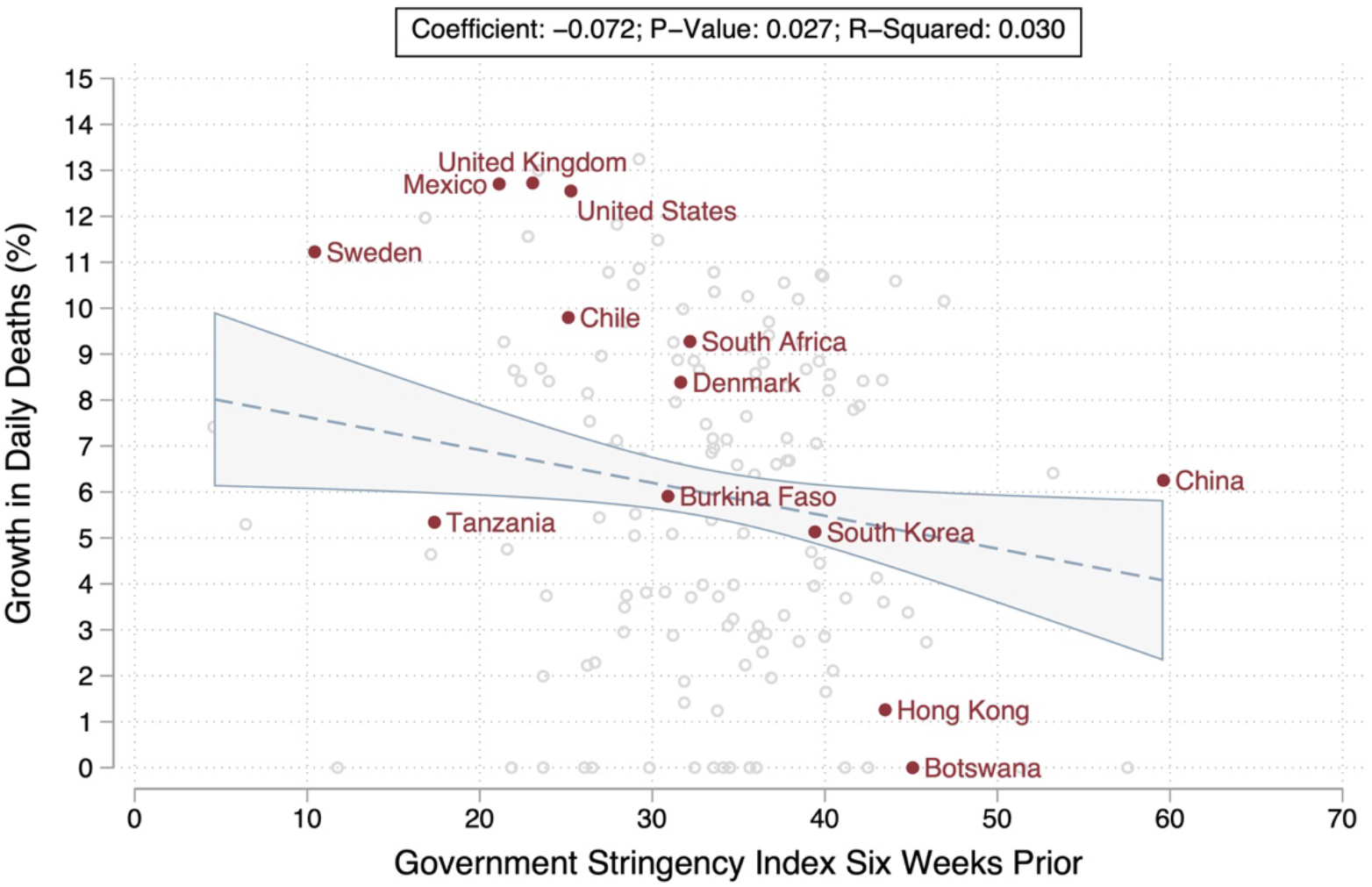
Degree of Government Response and Growth in Deaths Figure 3 depicts the relationship between the average degree of government response (measured on a scale of 0 to 100) and average growth rate in deaths by day, six weeks later. That is, the period over which the stringency of government response is averaged begins six weeks before the first case in a country, and runs up to six weeks before the endpoint of the analysis (May 27, 2020). In turn, the period over which the daily growth rate in deaths is averaged begins at the date of the first case and runs to May 27, 2020). Each additional point of stringency of government response is associated with a −0.072 (95% CI −0.134 to −0.008) reduction in the growth rate of daily deaths. A country with the maximum score (100) could expect to see an average daily death growth rate of 7.2% lower than a country with no measures in place. We average growth in deaths since the time of first case for each country to make comparisons on an equivalent time scale. Growth rates are computed as the first differences on a log scale. We also average government stringency six weeks prior to corresponding growth rates to capture likely downstream effects of government policies on later cases and deaths.

Finally, Figure 4 explores these relationships for six illustrative countries, comparing the government response (left y-axis) and growth rate in deaths (right y-axis) over time (normalized around days before and after a country’s first death). We observe that Botswana, which responded quickly and strongly, has almost no deaths and maintains this over time. South Korea, in turn, responds quickly despite being one of the first countries affected by the pandemic globally, and sees the second largest reduction in average daily growth of new deaths. Of note, South Korea also appears to sustain low death rates later in the epidemic after relaxing government restrictions, and also appears to benefit from a fast but relatively less stringent response. Colombia also sees an early response but is less fast that South Korea, although stronger, and in turn sees reductions in daily growth rates behind those of South Korea or Botswana, but quickly converges to low growth rates. Spain’s response only begins in earnest after the day of first death, and correspondingly takes longer to converge to slower growth rates. The United States, in turn, shows the slowest and weakest response and also the highest and most delayed path to epidemic control. The countries are broadly illustrative of patterns seen across 170 countries globally.

**Figure 4:**
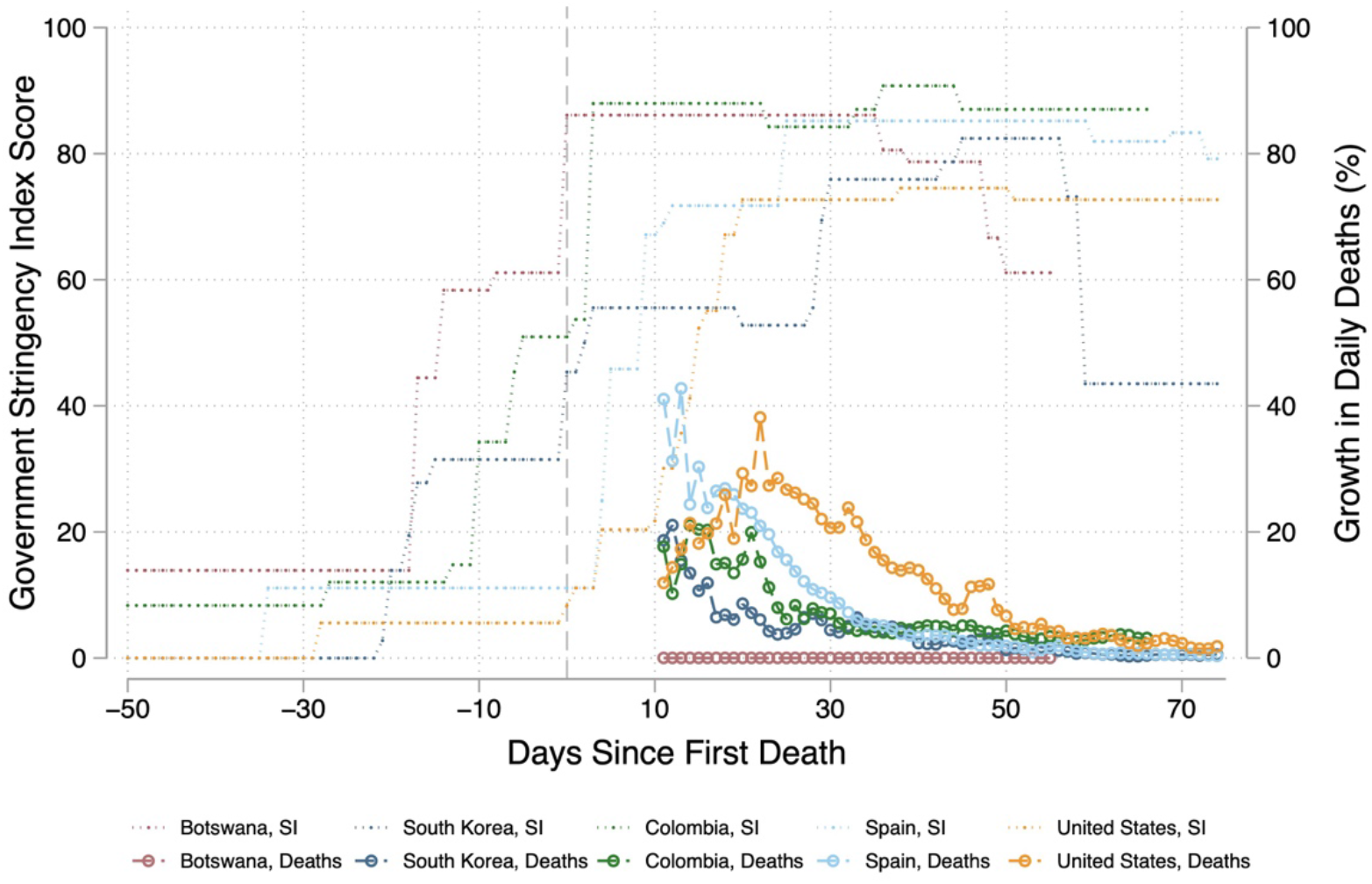
Government Response and Death by Day – Example Countries Figure 4 depicts five example countries to showcase the relationship between the strength and speed of government response and corresponding changes in the growth rate of daily deaths by day and country. The x-axis is normalized to days since first death in each country to make comparisons on an analogous time scale. The y-axes include the strength of the government response on a scale of 0 to 100 and average growth in new deaths per day since first case. Growth rates are computed as the first differences on a log scale. Countries that adopted more stringent measures, and did so more quickly, see lower death growth rates overall, and see the rate of growth fall more quickly.

### Regression results

Regression analysis allows us to further interrogate the descriptive relationships identified above. Table 1 presents the results for the cross-country models, which compare countries based on their performance over the whole period of analysis, as well as the time-series models, which compare “country-days” both across countries and across time. The former look at the effect of both speed and degree of government response (the latter averaged across the period excluding the most recent six weeks, similar to the lag in time series models) on the average daily growth rate in deaths, capturing variation between countries. The latter explore how the degree of government response six weeks prior relates to the growth rate in deaths in a given country on a given day. All models include controls for the time when a country experienced its first case and the magnitude of the first spike in cases. Further models are reported in the supplement Tables S3 and S4. Overall, the results strongly indicate that earlier and more stringent responses led to fewer deaths.

**Table 1:** Relationship Between Speed and Strength of Government Response and Deaths

|  | Difference in growth in deaths per day or unit change in government stringency index (95% CI)<sup>a</sup> | P-Value | N<sup>d</sup> |
| --- | --- | --- | --- |
| Cross Country Analysis<sup>b</sup> |  |  |  |
| Days until reached stringency index 40 since first case | 0.087 (0.056 to 0.118) | < 0.001 | 140 |
| Average government stringency six weeks prior | -0.080 (-0.121 to -0.039) | < 0.001 | 143 |
| Within and Across Country Analysis<sup>c</sup> |  |  |  |
| Government stringency six weeks prior | -0.039 (-0.069 to -0.008) | 0.014 | 3361 |
| Government stringency six weeks prior, adjusting for testing policy | -0.041 (-0.072 to -0.010) | 0.011 | 3640 |

<sup>a</sup> Growth in new deaths per day computed as the log difference and averaged since the day of the first case in each country.
<sup>b</sup> Analyses in this section control for the number of days since the first case in a country, the level of initial cases, and country-specific factors including the dependency ratio to capture the age structure of a given country, population density, health capacity in terms of beds and physicians per capita, and GDP per capita. N = number of countries included in analysis.
<sup>c</sup> Analyses in this section adjust for time and country fixed effects as well as the number of days since first case in a country and the level of initial cases.
<sup>d</sup> Observation in the cross-country analysis refer to countries. In the within and across country analysis, observations refer to country-days.

The first half of Table 1 shows how the speed and degree of government response relate to the average growth rate in deaths, including country-specific controls (the dependency ratio to capture the age structure of a given country, population density, health capacity in terms of beds and physicians per capita, and GDP per capita) that may substitute for or mediate the effect of government response measures. The results show that each additional day of delay increases the average growth rate in deaths by 0.087 percentage points (95% CI 0.056 to 0.118, P<0.001), while each additional SI point reduces the growth rate in deaths by 0.080 percentage points (95% CI −0.121 to −0.039, P<0.001). These estimates suggest a sizeable effect because even small differences in daily growth rates lead to large cumulative differences over months. For example, a week delay in reaching SI 40 would result in an average daily growth rate over half (0.609) a percentage point higher; over 90 days, that difference would lead to 1.7 times as many deaths overall.

The second half of Table 1 complements the cross-section analyses of countries with time-series analysis that compares country-days. Here we consider how the degree of country response affects the rate of growth in deaths six weeks later. The results indicate that higher stringency in the past leads to a lower growth rate in the present, with each additional point of stringency corresponding to a −0.039 percentage point reduction (95% CI −0.069 to −0.008, *P*=0.01). This means that a 10-point difference in SI would be expected to lead, six weeks later, to a daily growth rate in deaths nearly half a percentage point lower. Sustained over three months, this would correspond to a cumulative number of deaths 30% lower. The final row of Table 1 repeats these results with an additional control for countries’ testing policies, finding consistent results.

## Discussion

Our data show that speed and degree of government responses do indeed have a statistically robust and substantively significant relationship with deaths related to COVID-19. This study provides an early, global estimate of the effects of government NPIs.

Our study has several limitations. Like any policy intervention, the effect of the responses we measured is likely to be highly contingent on local political and social contexts. For instance, the state-by-state level response in the United States has been heterogeneous, and our data track responses only at the national level. Nor do we measure the extent to which government interventions are successfully implemented. In addition, the effects reported do not account for potential confounders that might have otherwise reduced deaths, such as seasonality and climate. While these factors have not yet been established for COVID-19, if they are, they will need to be accounted for to more reliably estimate the effect of government policies on growth in deaths. In spite of these limitations, our approach offers a global and comprehensive view of governmental response to COVID-19 to date with the best information available. By measuring a range of indicators, composite indices mitigate the possibility that any one indicator may be over- or mis-interpreted. By the same token, composite measures also make strong assumptions about what kinds of information are included. If the information left out is systematically correlated with the outcomes of interest, or systematically under- or overvalued compared to other indicators, such composite indices may introduce measurement bias.

Our data are in line with findings of the effects of similar NPIs on previous pandemics. Prior researchers have identified that earlier stringency measures decreased death rates in the 1918-1919 influenza pandemic by as much as 50%.^35,39^ An analysis of the 2009 influenza pandemic identified that variability in government stringency may have been the single biggest factor in determining country-to-country variability in disease impact.^40^ Regarding COVID-19, early data from China also support our findings that increased stringency decreases the transmission of SARS-CoV-2.^21,22,41,42^ An analysis of five policy categories on COVID-19 deaths in 11 European countries found a significant impact of interventions implemented several weeks before late March, though these results were strongly driven by the experiences of Spain and Italy.^19,43^ This study therefore concludes that for most European countries in late March it remained “too early to be certain that recent interventions have been effective.” Another impact assessment limited to Chinese locations outside Hubei, estimated reductions in R0 after control measures were introduced on January 23, 2020.^23^ A number of commentaries in medical journals have noted the apparent success of social distancing policies in China.^2,18,20,44^ Recent research has also analysed the association of public health interventions with hospitalization in the United States.^45^ Another study analysed the effects of 1,717 non-pharmaceutical interventions deployed in six countries and found that anti-contagion policies have prevented or delayed infections.^26^ Our data now provide the most comprehensive test of these ideas to date.

Going forward, it will be important to continue monitoring government responses as the pandemic evolves. More granular analyses looking at the implementation and effectiveness of national policies, the role of individual measures and various combinations of policies, as well as the role of subnational governments or other social institutions will be important. The fact that most governments adopted most measures makes it difficult to ascertain via regression analysis which specific government responses had more or less individual or combined effect on the spread of the disease. As countries start to ease their policies and experiment with different settings and mixtures of policies, it may be possible to assess the contribution of each NPI to the observed outcome. This is an important avenue for future research.

## Data Availability

All underlying data are freely available, and continuously updated, on the website of the Oxford COVID-19 Government Response Tracker: https://www.bsg.ox.ac.uk/research/research-projects/coronavirus-government-response-tracker

## Acknowledgements

We are grateful to the strong support from students and staff at the Blavatnik School of Government and across Oxford University for contributing time and energy to data collection. We thank Rafael Goldszmidt, Andy Eggers, and Devika Singh for helpful comments.

## References

1. World Health Organization. Coronavirus Disease (COVID-2019) Situation Reports, 1 June 2020. https://www.who.int/docs/default-source/coronaviruse/situation-reports/20200601-covid-19-sitrep-133.pdf?sfvrsn=9a56f2ac_4.

2. Wu Z, McGoogan JM. Characteristics of and Important Lessons From the Coronavirus Disease 2019 (COVID-19) Outbreak in China: Summary of a Report of 72 314 Cases From the Chinese Center for Disease Control and Prevention. JAMA. 2020;323(13):1239. doi:10.1001/jama.2020.2648.

3. Lurie N, Saville M, Hatchett R, Halton J. Developing Covid-19 Vaccines at Pandemic Speed. N Engl J Med. Published online March 30, 2020. doi:10.1056/NEJMp2005630.

4. Beigel JH, Tomashek KM, Dodd LE, et al. Remdesivir for the Treatment of Covid-19 — Preliminary Report. N Engl J Med. Published online May 22, 2020:NEJMoa2007764. doi:10.1056/NEJMoa2007764.

5. Hunter DJ. Covid-19 and the Stiff Upper Lip — The Pandemic Response in the United Kingdom. N Engl J Med. 2020;382(16):e31. doi:10.1056/NEJMp2005755.

6. Gates B. Responding to Covid-19 — A Once-in-a-Century Pandemic? N Engl J Med. 2020;382(18):1677–1679. doi:10.1056/NEJMp2003762.

7. Gostin LO, Wiley LF. Governmental Public Health Powers During the COVID-19 Pandemic: Stay-at-home Orders, Business Closures, and Travel Restrictions. JAMA. Published online April 2, 2020. doi:10.1001/jama.2020.5460.

8. Cowling BJ, Aiello AE. Public Health Measures to Slow Community Spread of Coronavirus Disease 2019. J Infect Dis. Published online March 20, 2020. doi:10.1093/infdis/jiaa123.

9. Fong MW, Gao H, Wong JY, et al. Nonpharmaceutical Measures for Pandemic Influenza in Nonhealthcare Settings—Social Distancing Measures. Emerg Infect Dis. 2020;26(5). doi:10.3201/eid2605.190995.

10. Haushofer J, Metcalf CJE. Which interventions work best in a pandemic? Science. Published online May 21, 2020:eabb6144. doi:10.1126/science.abb6144.

11. Viner RM, Russell SJ, Croker H, et al. School closure and management practices during coronavirus outbreaks including COVID-19: a rapid systematic review. Lancet Child Adolesc Health. 2020;4(5):397–404. doi:10.1016/S2352-4642(20)30095-X.

12. Wang CJ, Ng CY, Brook RH. Response to COVID-19 in Taiwan: Big Data Analytics, New Technology, and Proactive Testing. JAMA. 2020;323(14):1341. doi:10.1001/jama.2020.3151.

13. Steinbrook R. Contact Tracing, Testing, and Control of COVID-19—Learning From Taiwan. JAMA Intern Med. Published online May 1, 2020. doi:10.1001/jamainternmed.2020.2072.

14. Chu DK, Akl EA, Duda S, et al. Physical distancing, face masks, and eye protection to prevent person-to-person transmission of SARS-CoV-2 and COVID-19: a systematic review and meta-analysis. The Lancet. Published online June 2020:S0140673620311429. doi:10.1016/S0140-6736(20)31142-9.

15. Fauci AS, Lane HC, Redfield RR. Covid-19 — Navigating the Uncharted. N Engl J Med. 2020;382(13):1268–1269. doi:10.1056/NEJMe2002387.

16. Maxwell DN, Perl TM, Cutrell JB. “The Art of War” in the Era of Coronavirus Disease 2019 (COVID-19). Clin Infect Dis. Published online March 4, 2020. doi:10.1093/cid/ciaa229.

17. Esposito S, Principi N. School Closure During the Coronavirus Disease 2019 (COVID-19) Pandemic: An Effective Intervention at the Global Level? JAMA Pediatr. Published online May 13, 2020. doi:10.1001/jamapediatrics.2020.1892.

18. Anderson RM, Heesterbeek H, Klinkenberg D, Hollingsworth TD. How will country-based mitigation measures influence the course of the COVID-19 epidemic? The Lancet. 2020;395(10228):931–934. doi:10.1016/S0140-6736(20)30567-5.

19. Flaxman S, Gandy A, Unwin HJT, et al. Estimating the effects of non-pharmaceutical interventions on COVID-19 in Europe. Nature. Published online June 8, 2020. doi:10.1038/s41586-020-2405-7.

20. Kupferschmidt K, Cohen J. Can China’s COVID-19 strategy work elsewhere? Science. 2020;367(6482):1061–1062. doi:10.1126/science.367.6482.1061.

21. Chinazzi M, Davis JT, Ajelli M, et al. The effect of travel restrictions on the spread of the 2019 novel coronavirus (COVID-19) outbreak. Science. Published online March 6, 2020. doi:10.1126/science.aba9757.

22. Pan A, Liu L, Wang C, et al. Association of Public Health Interventions With the Epidemiology of the COVID-19 Outbreak in Wuhan, China. JAMA. Published online April 10, 2020. doi:10.1001/jama.2020.6130.

23. Leung K, Wu JT, Liu D, Leung GM. First-wave COVID-19 transmissibility and severity in China outside Hubei after control measures, and second-wave scenario planning: a modelling impact assessment. The Lancet. Published online April 8, 2020. doi:10.1016/S0140-6736(20)30746-7.

24. Davies NG, Kucharski AJ, Eggo RM, et al. Effects of non-pharmaceutical interventions on COVID-19 cases, deaths, and demand for hospital services in the UK: a modelling study. Lancet Public Health. Published online June 2020:S246826672030133X. doi:10.1016/S2468-2667(20)30133-X.

25. MacIntyre CR, Wang Q. Physical distancing, face masks, and eye protection for prevention of COVID-19. The Lancet. Published online June 2020:S0140673620311831. doi:10.1016/S0140-6736(20)31183-1.

26. Hsiang S, Allen D, Annan-Phan S, et al. The effect of large-scale anti-contagion policies on the COVID-19 pandemic. Nature. Published online June 8, 2020. doi:10.1038/s41586-020-2404-8.

27. Greco S, Ishizaka A, Tasiou M, Torrisi G. On the Methodological Framework of Composite Indices: A Review of the Issues of Weighting, Aggregation, and Robustness. Soc Indic Res. 2019;141(1):61–94. doi:10.1007/s11205-017-1832-9.

28. Alwin DF. The Use of Factor Analysis in the Construction of Linear Composites in Social Research. Sociol Methods Res. 1973;2(2):191–212. doi:10.1177/004912417300200202.

29. European Centre for Disease Prevention and Control (ECDC). Data on the Geographic Distribution of COVID-19 Cases Worldwide, 2020. https://www.ecdc.europa.eu/en/publications-data/download-todays-data-geographic-distribution-covid-19-cases-worldwide.

30. Dong E, Du H, Gardner L. An interactive web-based dashboard to track COVID-19 in real time. Lancet Infect Dis. Published online February 19, 2020. doi:10.1016/S1473-3099(20)30120-1.

31. Lipsitch M, Swerdlow DL, Finelli L. Defining the Epidemiology of Covid-19 — Studies Needed. N Engl J Med. 2020;382(13):1194–1196. doi:10.1056/NEJMp2002125.

32. Wallinga J, Teunis P. Different Epidemic Curves for Severe Acute Respiratory Syndrome Reveal Similar Impacts of Control Measures. Am J Epidemiol. 2004;160(6):509–516. doi:10.1093/aje/kwh255.

33. Atkins KE, Wenzel NS, Ndeffo-Mbah M, Altice FL, Townsend JP, Galvani AP. Under-reporting and case fatality estimates for emerging epidemics. BMJ. 2015;350(mar163):h1115–h1115. doi:10.1136/bmj.h1115.

34. Zou L, Ruan F, Huang M, et al. SARS-CoV-2 Viral Load in Upper Respiratory Specimens of Infected Patients. N Engl J Med. 2020;382(12):1177–1179. doi:10.1056/NEJMc2001737.

35. Bootsma MCJ, Ferguson NM. The effect of public health measures on the 1918 influenza pandemic in U.S. cities. Proc Natl Acad Sci. 2007;104(18):7588–7593. doi:10.1073/pnas.0611071104.

36. Dieleman JL, Templin T. Random-Effects, Fixed-Effects and the within-between Specification for Clustered Data in Observational Health Studies: A Simulation Study. Dalby AR, ed. PLoS One. 2014;9(10):e110257. doi:10.1371/journal.pone.0110257.

37. Leszczensky L, Wolbring T. How to Deal With Reverse Causality Using Panel Data? Recommendations for Researchers Based on a Simulation Study. Sociol Methods Res. Published online November 14, 2019. doi:10.1177/0049124119882473.

38. Hatchett RJ, Mecher CE, Lipsitch M. Public health interventions and epidemic intensity during the 1918 influenza pandemic. Proc Natl Acad Sci. 2007;104(18):7582–7587. doi:10.1073/pnas.0610941104.

39. Markel H, Lipman HB, Navarro JA, et al. Nonpharmaceutical Interventions Implemented by US Cities During the 1918-1919 Influenza Pandemic. JAMA. 2007;298(6):644. doi:10.1001/jama.298.6.644.

40. Cauchemez S, Van Kerkhove MD, Archer BN, et al. School closures during the 2009 influenza pandemic: national and local experiences. BMC Infect Dis. 2014;14(1):207. doi:10.1186/1471-2334-14-207.

41. Tian H, Liu Y, Li Y, et al. An investigation of transmission control measures during the first 50 days of the COVID-19 epidemic in China. Science. Published online March 31, 2020. doi:10.1126/science.abb6105.

42. Ji T, Chen H-L, Xu J, et al. Lockdown contained the spread of 2019 novel coronavirus disease in Huangshi city, China: Early epidemiological findings. Clin Infect Dis. Published online April 7, 2020. doi:10.1093/cid/ciaa390.

43. Flaxman S, Mishra S, Gandy A, et al. Report 13: Estimating the Number of Infections and the Impact of Non-Pharmaceutical Interventions on COVID-19 in 11 European Countries. Imperial College London; 2020. doi:10.25561/77731.

44. Inglesby TV. Public Health Measures and the Reproduction Number of SARS-CoV-2. JAMA. 2020;323(21):2186. doi:10.1001/jama.2020.7878.

45. Sen S, Karaca-Mandic P, Georgiou A. Association of Stay-at-Home Orders With COVID-19 Hospitalizations in 4 States. JAMA. Published online May 27, 2020. doi:10.1001/jama.2020.9176

